# A variant prioritization tool leveraging multiple instance learning for rare Mendelian disease genomic testing

**DOI:** 10.1101/2024.04.18.24305632

**Authors:** Ho Heon Kim, Ju Yeop Baek, Heonjong Han, Won Chan Jeong, Dong-Wook Kim, Kisang Kwon, Yongjun Song, Hane Lee, Go Hun Seo, Jungsul Lee, Kyoungyeul Lee

**Affiliations:** Research and Development Center, 3billion, Inc., 416 Teheran-ro, 06193 Seoul, Republic of Korea

## Abstract

**Background:** Genomic testing such as exome sequencing and genome sequencing is being widely utilized for diagnosing rare Mendelian disorders. Because of a large number of variants identified by these tests, interpreting the final list of variants and identifying the disease-causing variant even after filtering out likely benign variants could be labor-intensive and time-consuming. It becomes even more burdensome when various variant types such as structural variants need to be considered simultaneously with small variants. One way to accelerate the interpretation process is to have all variants accurately prioritized so that the most likely diagnostic variant(s) are clearly distinguished from the rest.

**Methods:** To comprehensively predict the genomic test results, we developed a deep learning based variant prioritization system that leverages multiple instance learning and feeds multiple variant types for variant prioritization. We additionally adopted learning to rank (LTR) for optimal prioritization. We retrospectively developed and validated the model with 5-fold cross-validation in 23,115 patients with suspected rare diseases who underwent whole exome sequencing. Furthermore, we conducted the ablation test to confirm the effectiveness of LTR and the importance of permutational features for model interpretation. We also compared the prioritization performance to publicly available variant prioritization tools.

**Results:** The model showed an average AUROC of 0.92 for the genomic test results. Further, the model had a hit rate of 96.8% at 5 when prioritizing single nucleotide variants (SNVs)/small insertions and deletions (INDELs) and copy number variants (CNVs) together, and a hit rate of 95.0% at 5 when prioritizing CNVs alone. Our model outperformed publicly available variant prioritization tools for SNV/INDEL only. In addition, the ablation test showed that the model using LTR significantly outperformed the baseline model that does not use LTR in variant prioritization (*p*=0.007).

**Conclusion:** A deep learning model leveraging multiple instance learning precisely predicted genetic testing conclusion while prioritizing multiple types of variants. This model is expected to accelerate the variant interpretation process in finding the disease-causing variants more quickly for rare genetic diseases.

## Introduction

Of the ~10,000 rare diseases affecting ~400 million people globally, more than 80% are reported to have a genetic cause^1^. With the advent of next-generation sequencing (NGS), not only have genomic tests become readily accessible for diagnosing patients with suspected rare genetic diseases, but also vast number of novel rare variants are being discovered and shared worldwide, becoming a rich resource for predicting variant pathogenicity ^2–4^. The more variant data accumulate, the more precise variant classification will be, resulting in more accurate molecular diagnosis and higher diagnostic yield. However, each variant needs to be carefully curated before it becomes either diagnostic or disregarded or even simply being shared with the community, and as a large number of variants are flooded out each day, a systematic approach to curate variants is essential.

When a genomic testing is performed on a patient with a suspected rare disease, a total of ~100K variants and ~5 million variants are identified by exome and genome sequencing, respectively ^5,6^. Most of these variants are common variants that are unlikely to be causal for rare diseases, and therefore they are filtered out to leave ~5% of the variants ^6^. Once various evidence and rules are applied to each variant according to the American College of Medical Genetics and Genomics (ACMG)-recommended variant classification guidelines, typically 50~100 variants predicted to change the protein consequence within known rare disease genes are left for the clinical/medical geneticists to manually curate each variant in the context of the patient’s clinical symptoms. Although significantly smaller than the initial number of variants called, manually curating 50~100 variants to find 1~2 disease-causing variants is a labor-intensive and time-consuming task.

Each laboratory must use a series of tools or rules beyond the ACMG guidelines to prioritize the variants as automatically as possible. For example, Bayesian score can be calculated based on the ACMG rules added to each variant ^7^. Variants may be scored according to the degree of symptom similarities between the phenotype of the disease that the variant may cause and the patient’s clinical symptoms. These scores and evidence-based rules then need to be somehow combined and used to prioritize the variants in the order of how likely they can explain the patient’s clinical symptoms. There have been multiple attempts to develop variant prioritization (VP) tools based on machine learning algorithms ^2,8–12^. However, even though these VP tools have shown potential to accelerate the variant interpretation process, there are obvious limitations. First, to our knowledge, none of the publicly available tools can simultaneously prioritize different types of variants such as single nucleotide variants (SNV), small insertions and deletions (INDEL), and structural variants (SV) including copy number variants (CNV). Numerous studies show that CNVs are the cause of approximately 10% of rare genetic diseases^13,14^. Second, other VP tools only rank the variants from more likely pathogenic to less likely pathogenic without considering how well the variant could explain the patient’s phenotype. Therefore, the top-1 variant in a positive case will have the same rank score as the top-1 variant in a negative case. Predicting the genetic testing results accelerates variant interpretation without exploring candidate variants.

The previous version of our tool, 3ASC v1 used Random Forest (RF), a machine learning algorithm based on ensembles of trees based on ACMG/AMP guidelines^11^. One of the key and superior features of 3ASC v1 was considering variant quality in prioritization by incorporating adjusted scores for predicted false-positive variants. Here, we introduce 3ASC v2 that outperforms publicly available VPs by using multiple instance learning (MIL) and learning to rank (LTR) to simultaneously prioritize SNV/INDEL and CNV.

## Methods

### Problem definition and multiple instance learning

MIL is a weakly supervised learning model for which the learner feeds a single class label assigned to a bag of instances, instead of receiving a set of instances that are individually labeled^15^. In the *standard* multiple instance assumption, each instance has a hidden class label as either a positive or a negative label, and a bag is deemed positive only if it contains at least one positive instance^15,16^. A model levering MIL is typically designed to predict the bag label^16^. MIL also has a characteristic called *permutation invariance*, which refers to conditions where the output value remains unchanged even if the order of the input data changes. Since both the standard MIL assumption and permutation invariance are applicable to digital pathology, MIL is often used to predict the malignancy, which is considered the test result, from a whole slide image ^17,18^.

Variant interpretation is a process that involves reviewing each variant, regardless of its type, to evaluate all the annotated evidence and determine the classification for each variant accordingly ^19^. When a patient has a pathogenic or likely pathogenic (P/LP) variant in a gene-disease that could explain the patient’s given symptoms well, the variant is reported, and the genomic test result is considered diagnostic or positive. Inspired by the MIL based approach, our problem addresses the genomic test results as the bag label, and the causal variant as the instance label.

In contrast to the *standard* MIL assumption, we considered the instance label for each variant to be either causal or noncausal. Additionally, to predict these instance labels, we utilized attention mechanism-based MIL to prioritize candidate causal variants.

### Variant prioritization leveraging MIL: 3ASC

The MIL model, which we named 3ASC, was constructed to incorporate multiple types of variants and to prioritize potentially causal variants (Figure 1). Specifically, the model was designed to incorporate both SNV/INDEL and CNV as multiple inputs. We then embedded these input variant features into a low-dimensional space using neural network encoders. To address the different dimensionality of SNV/INDEL and CNV features, distinct encoders were constructed for each type of variant. Then, we additionally attached the shared embedding layer from SNV/INDEL and CNV encoders. This transformation projected the encoded features onto the manifold, where they could be on semantically equivalent dimensions. Since it is essential to aggregate these embedded instance vectors to predict the genomic test results, the model has an attention-based pooling layer with attention weights derived from a gated attention layer^15^. Finally, we added the classifier to predict the bag label, the genomic test results. For VP, we reused the attention weights indicating the level of importance in predicting the genomic test results to classify the instance label. The attention-weighted instance embedding vectors were fed into the instance classifier.

**Figure 1.**
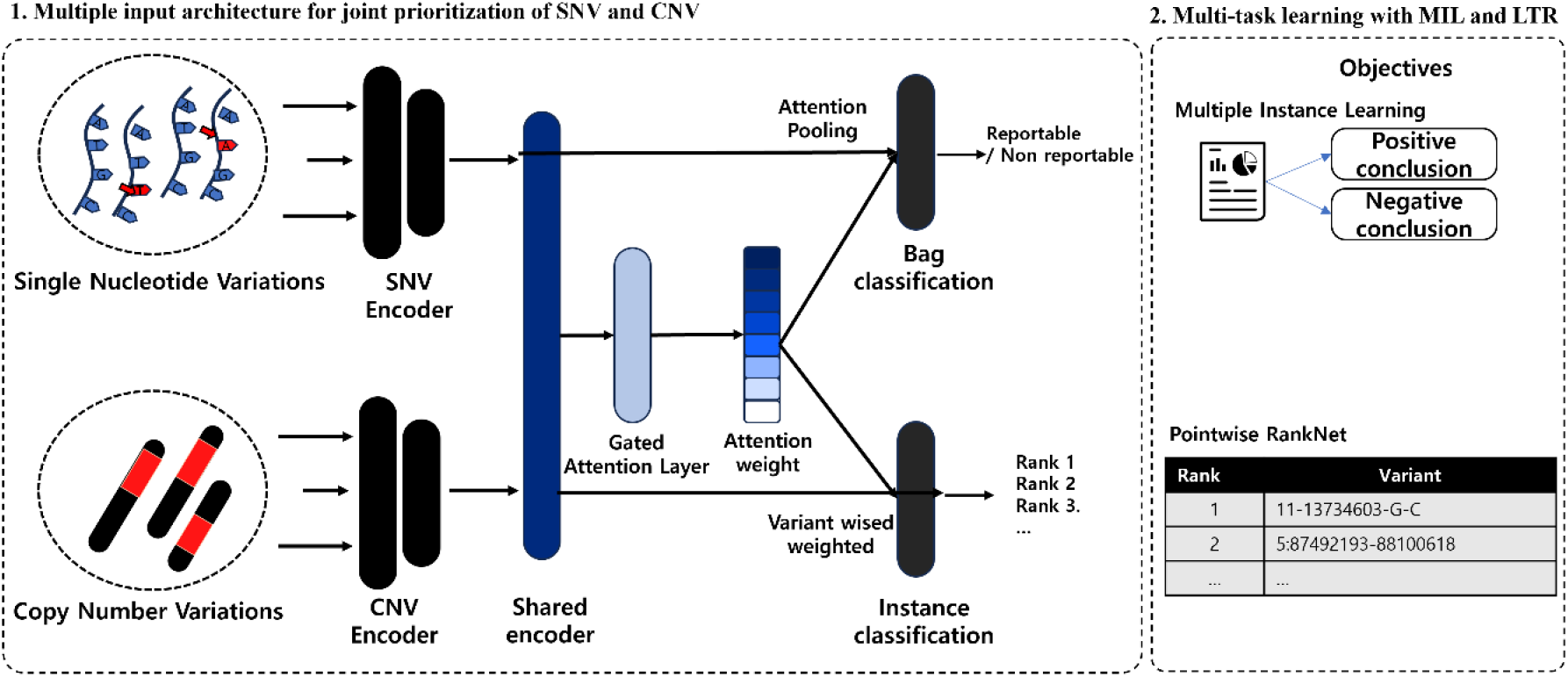
Overall architecture of 3ASC. A) 3ASC simultaneously feeds both SNV/INDEL and CNV, and each of the variant encoders compresses the variant information in a separate dimension. The shared encoder then embeds SNV/INDEL and CNV in a semantically shared dimension. On this dimension, the bag classifier predicts the genomic test result by using the attention weight and the instance classifier predicts the rank of all variants, regardless of its type. B) 3ASC performs two tasks simultaneously: predicting the genomic test result using MIL and ranking the variants using LTR.

### Pointwise RankNet

To achieve optimal VP, we additionally employed RankNet which is a differentiable loss function of deep learning during the training phase ^20,21^. Instead of predicting a confidence score of 1 for the causal variant when used in the classification loss function, RankNet enables a model to rank the causal variant higher than the non-causal variants even if the confidence score is not 1. This approach may be more suitable for VP, as the order of each variant is more important than its predictive value. We simplified the relevance level of RankNet to either 0 or 1, instead of a range from 1 to 5. Since it is not necessary to compare the ranks between non-causal variants, we have modified RankNet to focus solely on causal variants by comparing the ranks of the causal variants that are higher than those of the non-causal variants, we named pointwise RankNet. We implemented pointwise RankNet by defining *P*^*ij*^ as the learned probability that the rank of causal variants, *i*-th variant, should be higher than that of other non-causal *j*-th variants. For general rank comparison where both variants are causal variants, we used the proxy indicator *S*^*ij*^ ∈ {0, ±1} to calculate known probability 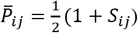 inspired by previous studies (Equation 1) ^21^. Then, the cross-entropy cost function was applied, and divided by the number of variants *n*.

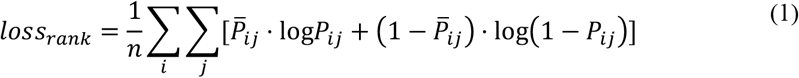

Finally, we formulated the loss function by averaging three of loss function including two binary cross-entropies for instance label, and for bag label, and pointwise RankNet.

### Patient exome sequencing data

This study used de-identified exome sequencing data from 23,115 patients with suspected rare disease, generated for diagnostic purposes at a single reference laboratory between Jan 01, 2021 to Sep 05, 2023. Patient samples were obtained as EDTA blood, buccal swabs, dried blood spots or extracted genomic DNA. All protein-coding regions of known human genes (~22,000) were captured by xGen Exome Research Panel v2 (Integrated DNA Technologies, Coralville, Iowa, USA) and sequenced with Novaseq6000 (Illumina, San Diego, CA, USA) as 150 bp paired-end reads. Sequencing reads were aligned to the human reference genome (GRCh37/hg19 from NCBI, February 2009) using BWA-MEM (v.0.7.17) ^22^. SNV/INDEL were called following the GATK best practices (GATK v3.8) and annotated using Ensembl Variant Effect Predictor (VEP v104) ^23,24^. CNV were called using CoNIFER and 3bCNV, an in-house CNV caller ^25^. Variants were then classified according to the ACMG/AMP guidelines using 3billion’s bioinformatics pipeline, EVIDENCE, as previously described ^26^. In brief, for SNV/ INDEL, 41 features including posterior probability of pathogenicity, semantic similarity between the patient’s phenotypes and the disease phenotypes (i.e. symptom similarity), and the variant quality were used for classification. For CNVs, 3 features were used, including the posterior probability of variant pathogenicity, the maximum symptom similarity among all known disease genes within the CNV interval, and the number of affected genes (Supplementary document 1). For each patient, variants with an allele frequency > 5% in gnomAD 2.0 or >1% internally were determined to be too common to cause a rare disease and filtered out ^27^. Variants reported as likely benign/benign and variants in genes that are not yet associated with a monogenic disorder in Online Mendelian Inheritance in Man (OMIM), Human Phenotype Ontology (HPO), Genomic OrphaNet, or Clinical Genomic Database (CGD) were also filtered out ^28–31^. The final list of variants was manually curated by the medical geneticists, and the variant that is likely to explain the patient’s symptoms was selected for reporting. The final reports were categorized as positive or negative depending on the presence of a pathogenic variant that can explain the patient’s phenotype.

### Ethics

The study was approved by the institutional review board at Korea National Institute for Bioethics Policy (P01-202308-02-001). Informed consent was determined unnecessary with the study only involving anonymous and de-identified retrospective data.

### Model validation

Model validation was performed as follows: First, we conducted a 5-fold cross-validation on the entire dataset to identify both area under the receiver operating characteristics (AUROC) for the genomic test results and the hit rate at *k* for VP. The hit rate at *k* is derived from averaging each patient’s hit at *k*, which refers to the existence of a relevant item within the predicted most relevant *k* items. It is considered that there is a hit if there is at least one causal variant among the top-*k* items. We also conducted an ablation test to assess the impact of the pointwise RankNet. The significance of the pointwise RankNet loss-to-train model was determined by using the Mann-Whitney U test. In addition, to evaluate the performance of predicting causalities for SNV/INDEL, we compared our model with two popular public VP tools, Exomiser and LIRICAL^2,32^. For Exomiser, “EXOMISER_VARIANT_SCORE” was used to prioritize variants. For LIRICAL, the ‘compositeLR’ score which is already used for ranking within the program was used^32^. We calculated the top-*k* hit rate for each model and observed that our model outperformed across all *k* values by a considerable margin. For model transparency, we employed the permutation feature importance to calculate the feature importance for both AUROC score and the top- *k* hit rate ^33^.

To assess the likelihood of type 1 statistical error, the effect sizes were calculated based on the type of variable and the number of groups compared. Given the risk for statistical power being inflated by comparing large population, which is almost always the case, we also calculated effect sizes to measure the magnitude of group differences in terms of means or proportions. In this study, all statistical tests were performed using a 2-sided *P* value threshold of <0.05. Effect sizes were interpreted as follows: Cramer V≈0.01: small; Cramer V≈0.30: moderate, and Cramer V≈0.50: large, Cohen d≈0.20 Cohen d≈0.50, Cohen d≈0.80). The model and experimental data are available at https://github.com/4pygmalion/ASC3.

## Results

### Overall characteristics

Of the 23,115 patients, 35.3% (n=8,157) received a positive report and 64.8% (n=14,998) received a negative report (Table 1). Although the proportion of male was significantly different (54.95% [4,483/8,157] vs 56.76% [8,514/14,998]; p-value: 0.008), the effect size was small (Cramer’s V=0.017). The difference in onset between the two groups was also statistically significant with 27.97% of positive samples being neonatal onset, while 33.6 % of negative samples being adult onset, but the effect size was small (p<.001; Cramer’s V=0.219). Finally, the difference in the number of phenotypes between the two groups was also significantly different but the effect size was small (p<.001; Cohen’s *D*=0.261). The majority of the patients had a causal variant that was SNV/INDEL (96.00%, 7,871/8,157) and only 4.80% (392/8,157) had a CNV. There were 66 patients reported with both an SNV/INDEL and a CNV. In both groups, the abnormality of the nervous system was the most common clinical indication (33.77% [3,559/8,157] vs 40.47% [5,627/14,998]).

**Table 1.**
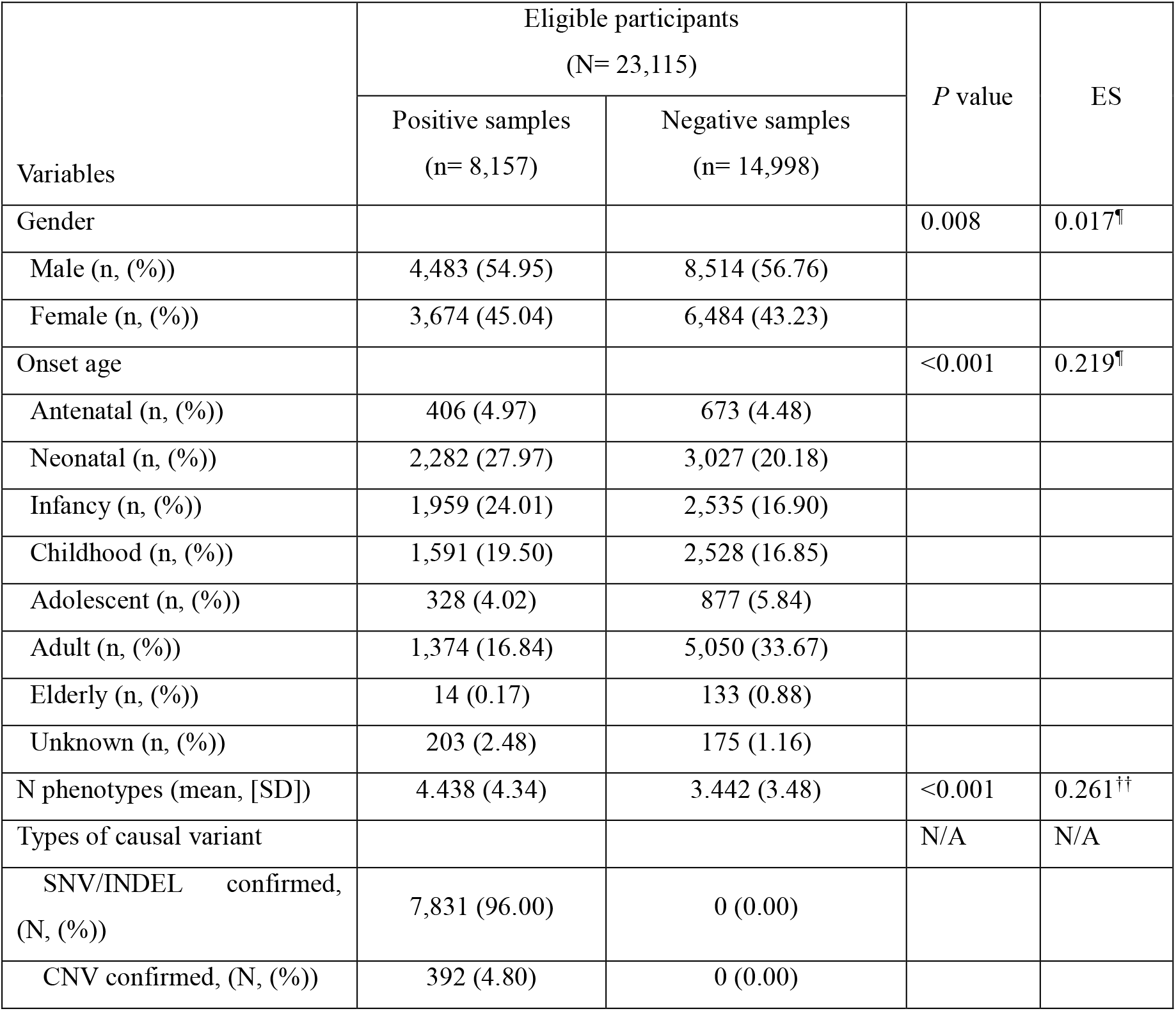
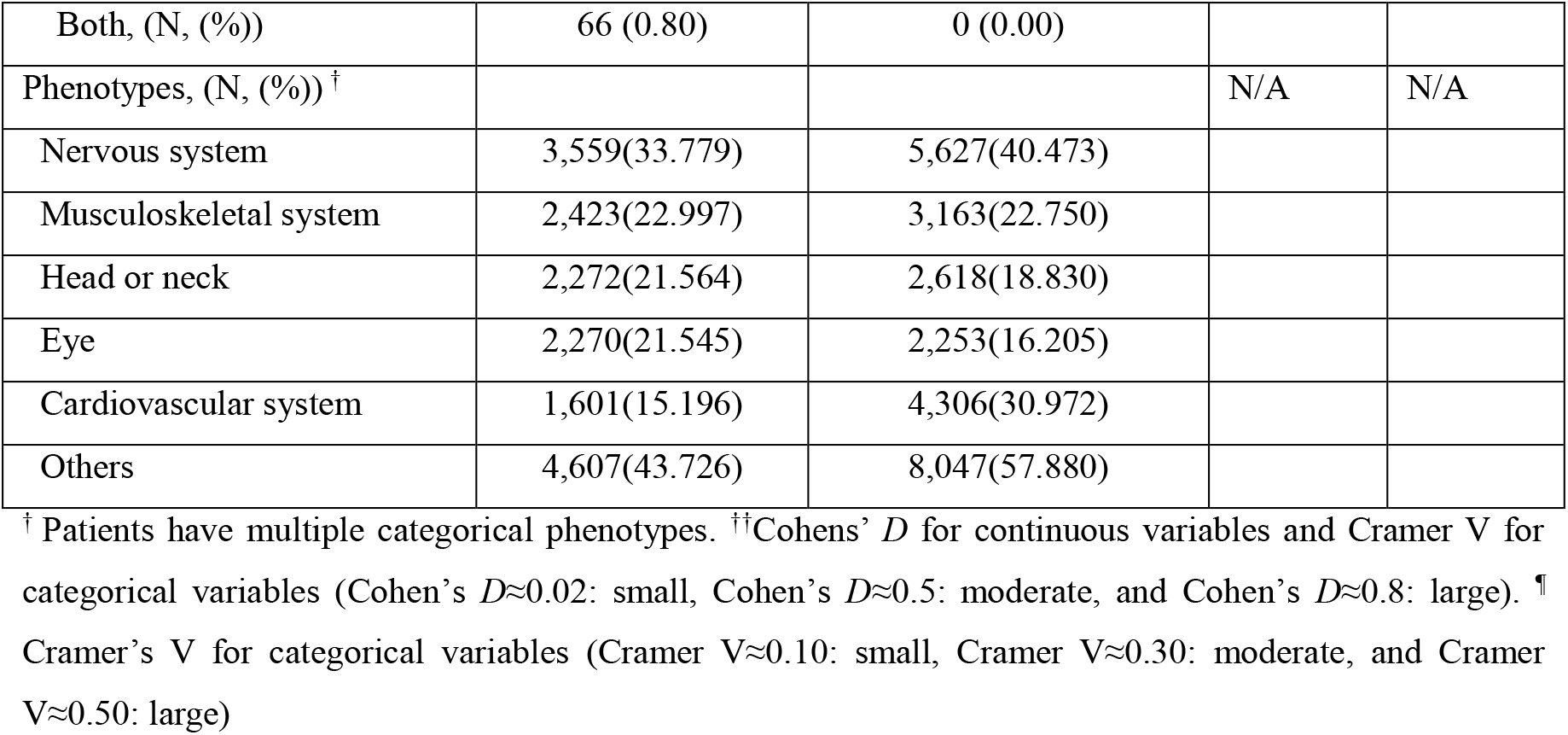
Demographic characteristics.

### Model performance

Through 5-fold cross-validation, the average AUROC for 3ASC predicting the genomic test results was calculated to be 0.92. For positive patients with one or more SNV/INDEL and CNV, the average hit rate for top-1 rank prediction was 73.9% with a standard deviation (SD) of 2.86%. For top-2 to 5, the average hit rate increased to 88.2% (SD: 1.61%), 93.1% (SD: 0.83%), 95.7% (SD: 0.36%), and 96.8% (SD: 0.26%). For positive patients diagnosed with a CNV, the average hit rate was 72.2% (SD: 6.38%) for top-1 rank prediction and 95.0% (SD: 5.06%) for top-5 (Figure 2).

**Figure 2.**
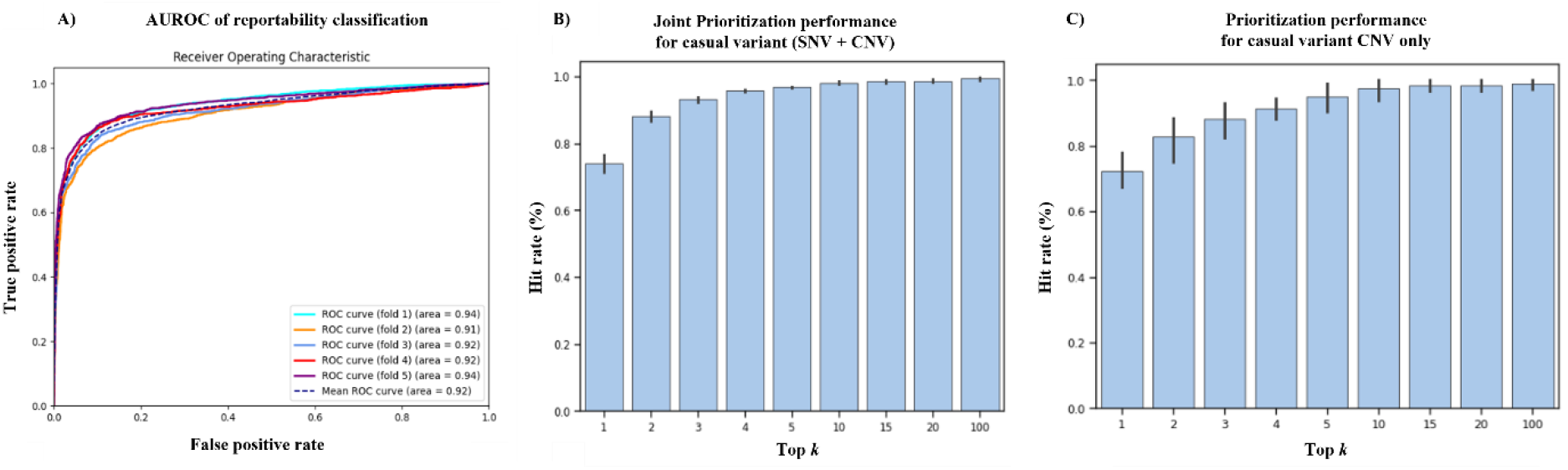
3ASC performance. A) AUROC for the genomic test results, B) Prioritization performance for causal variant in positive patients with either or both SNV/INDEL and CNV, C) Prioritization performance for causal variant in positive patients with CNV only.

### Ablation test: pointwise RankNet

The ablation test to compute the effectiveness of pointwise RankNet was conducted, and we trained two models by using pointwise RankNet and not using pointwise RankNet. For prioritizing causal SNV/INDEL or CNV, the model using pointwise RankNet significantly outperformed the model not using pointwise RankNet (*P*=0.007; respectively). The lower bound of the hit rate at top 5 for both SNV/INDEL and CNV from the model using pointwise RankNet was greater than the upper bound of the model not using pointwise RankNet (0.964 vs 0.960). Also, the deviation of hit rate at top 5 both SNV/INDELand CNV in the model using Pointwise RankNet was smaller than that of the model not using pointwise RankNet. Similarly, for ranking causal CNV, the model using pointwise RankNet significantly outperformed the model not using pointwise RankNet (p=0.007) (Figure 3).

**Figure 3.**
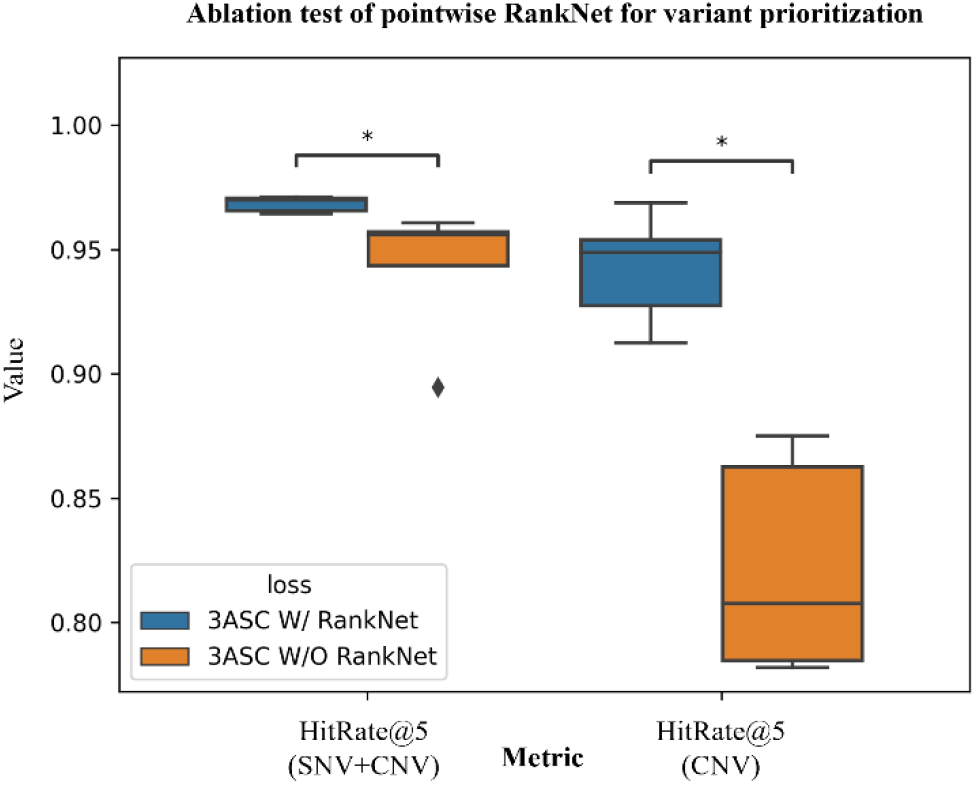
Ablation test of pointwise RankNet for VP

### SNV/INDEL prioritization benchmark

We compared 3ASC and publicly available tools, Exomiser, and LIRICAL as benchmarks and tested patients with causal SNV/INDEL only in test dataset (n=1,568). Our model outperformed both Exomiser and LIRICAL. The hit rates at top-1 and top-5 were 74.4% and 97.0% for 3ASC while they were 59.9% and 71.6% for Exomiser and 53.7% and 54.3% for LIRICAL, respectively (Figure 4).

**Figure 4.**
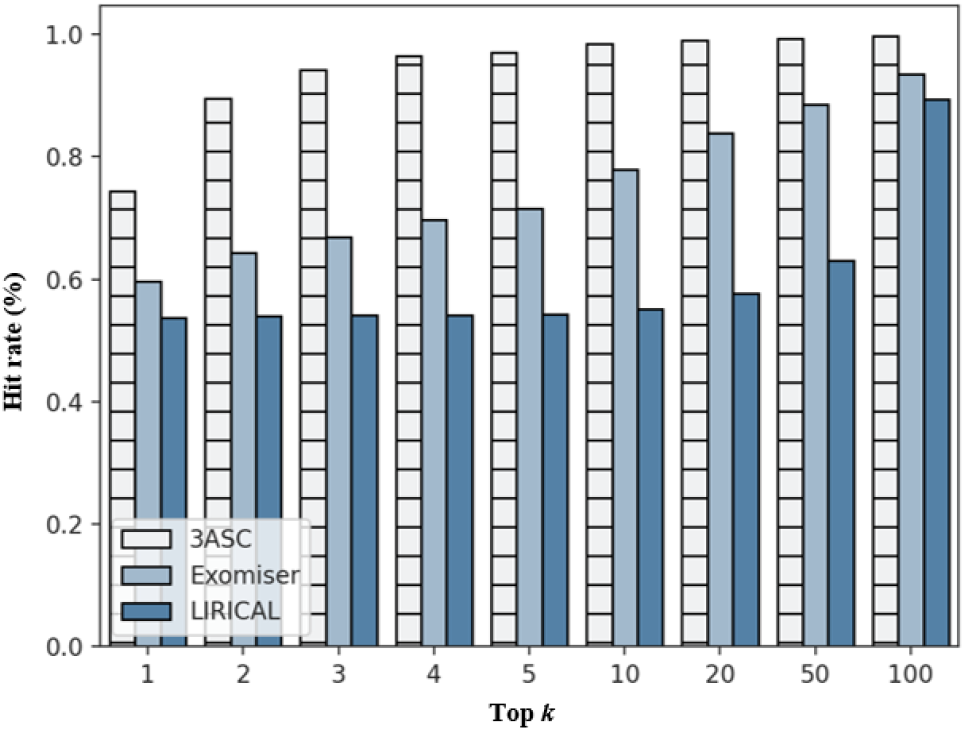
Benchmarking for SNV/INDEL prioritization.

### Post hoc interpretation

For model interpretation, permutation feature importance was conducted by shuffling specific feature values along rows while keeping other features fixed for both predicting the genomic test results and VP. For predicting the genomic test results, the decrease in AUROC was most pronounced when randomly symptom similarity was permuted (ΔAUROC=0.191). This was followed by variant allele frequency (VAF, 0.05), PVS1 rule (0.03) related to null variants that usually result in a loss of function, and matching inheritance between disease inheritance and patient genotype (0.02) (Figure 5-Figure 5A). For VP, the decrease in hit rate at 5 was also most pronounced for permutation of the symptom similarity (Δhit rate at 5=0.268, Figure 5-B). PVS1 (0.06), VAF (0.03), and matching inheritance (0.02) were the next features impacting the results.

**Figure 5.**
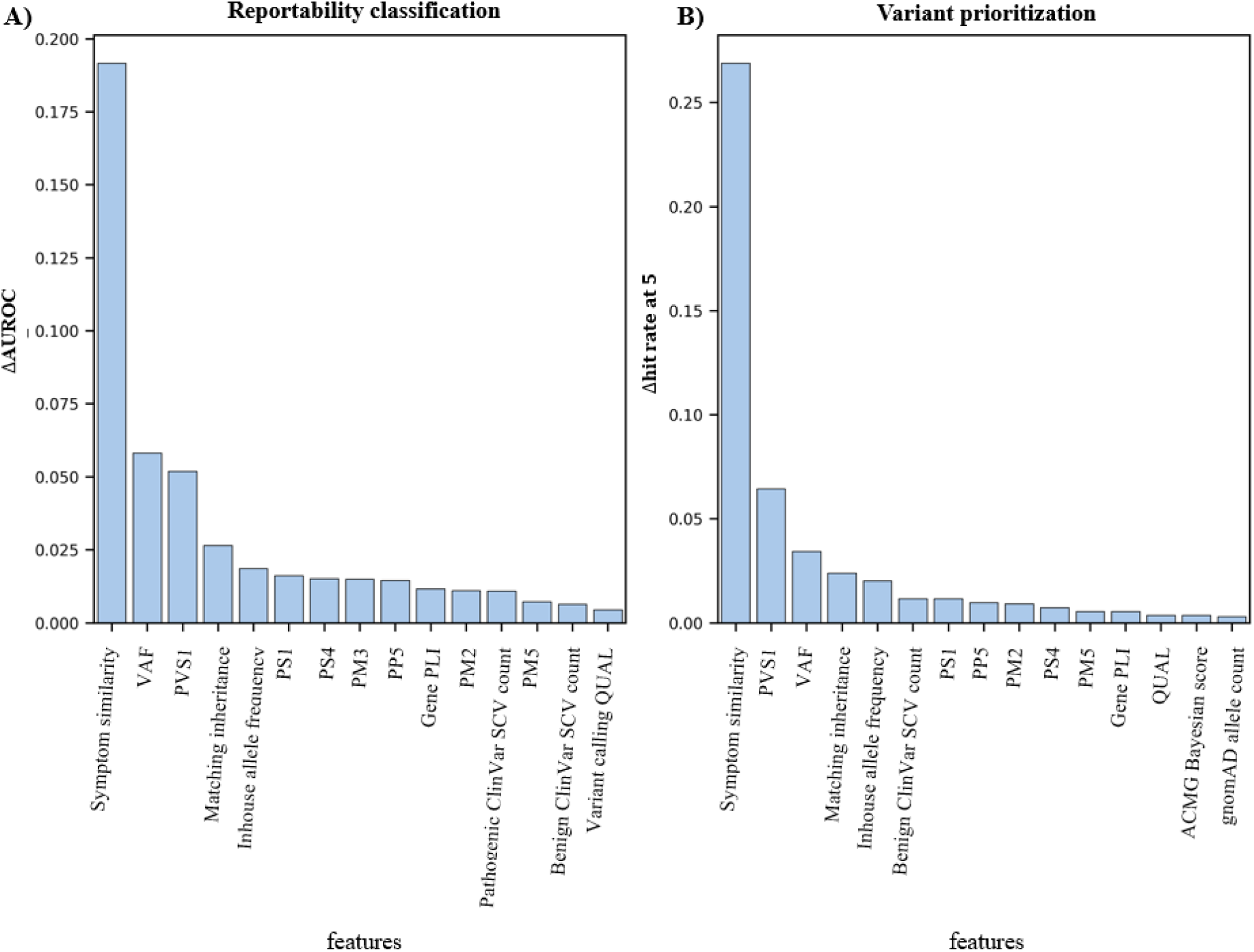
Top 15 feature importance of 3ASC. A) Difference in AUROC between the original data and the shuffled data for predicting the genomic test results, B) Difference in hit rate at 5 between the original data and the shuffled data for VP.

### Case examples

A male patient in the age range of 0-5 with global developmental delay, abnormal intestine morphology, facial dysmorphism, mild intellectual disability, and hearing loss had a dual diagnosis with a homozygous likely pathogenic NM_001080516.2:c.88C>T (NP_001073985.1:p.Arg30Ter) variant in *GRXCR2* associated with ‘Deafness, autosomal recessive 101 (OMIM: 615837)’ and a heterozygous pathogenic ~2.6Mb large deletion variant NC_000005.9:g.(?_113366)_(2,755,155_?)del in 5q14.3 associated with ‘Cri-Du-Chat syndrome (OMIM: 123450)’. 3ASC had ranked the deletion variant first (rank 1) with a model confidence score of 0.99, and the NM_001080516.2:c.88C>T variant third (rank 3) a model confidence score of 0.66. When all rare variants including the CNV projected by 3ASC embedding layer were plotted on the manifold, variants with high Bayesian scores and symptom similarity scores were clustered together. Furthermore, we could see that despite of the different features of each variant type, SNV/INDEL and CNV clustered semantically (Figure 6-A).

**Figure 6.**
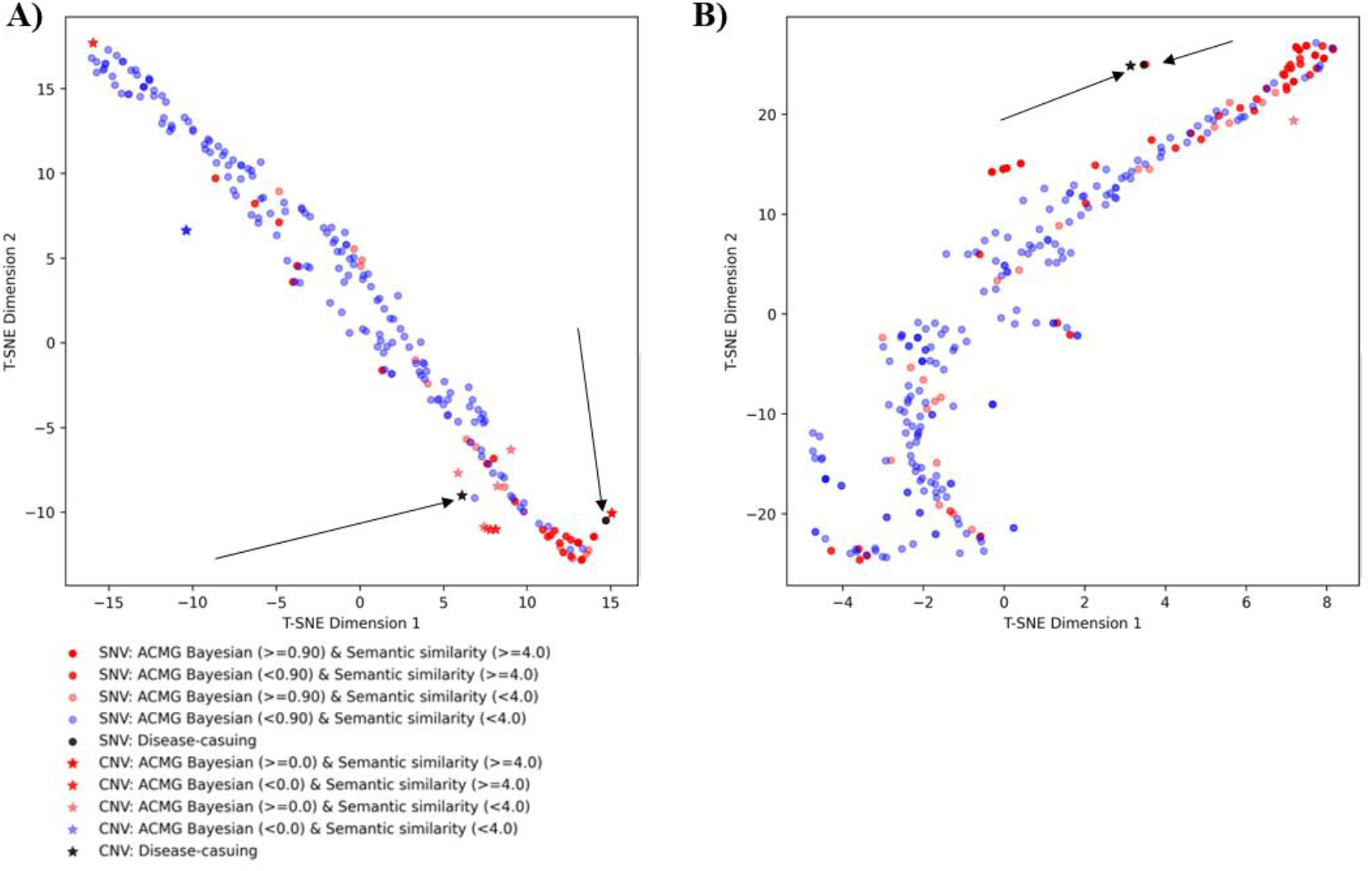
Multiple types of inputs (SNV/INDEL and CNV) encoded feature visualized on a manifold. Black arrows indicate causal variants. A) Variant manifold for the patient with a dual diagnosis of deafness and Cri-du-chat syndrome. B) Variant manifold for the patient with a dual diagnosis with an SNV and a CNV.

Another male patient in the age range of 0-5 with muscle weakness, strabismus, and striatal T2 hyperintensity on brain MRI had a dual diagnosis of ‘Neurodevelopmental disorder with hypotonia, stereotypic hand movements, and impaired language (OMIM: 613443)’ and ‘Convulsions, familial infantile, with paroxysmal choreoathetosis (OMIM: 602066)’. This patient had a heterozygous pathogenic NM_145239.3: c.649C>T (NP_660282.2:p.Arg217Ter) variant in the *PRRT2* gene, and a heterozygous ~101Kb deletion pathogenic variant NC_000005.9:g.(?_88,018,421)_(88,119,605_?)del(GRCh37) spanning the *MEF2C* gene. The NM_145239.3: c.649C>T variant was ranked third (rank 3) with a model confidence score of 0.36, and the CNV was ranked first (rank 1) with a model confidence score of 0.83. On the manifolds, the two variants semantically (Figure 6-B).

## Discussion

As genomic testing is becoming a routine diagnostic test for patients with suspected rare diseases, variant prioritization (VP) is also become essential in effectively managing large number of variants in each of a large number of test cases. The VP process becomes even more complicated when various variant types such as SNV/INDEL, CNV, repeat expansion variants, mobile element insertion variants need to be considered simultaneously as potential causal variants. Here, we developed a novel MIL-based model that can effectively prioritize rare variants with high AUROC and hit rate at 5. Our model attempts to use as many variables as possible that medical geneticists consider when interpreting variants as features. Such variables include symptom similarity score, Bayesian score for variant pathogenicity, and whether the variant has been reported as pathogenic elsewhere, etc. By integrating ACMG/AMP variant interpretation guideline into the deep learning model, our model learned the variant interpretation pattern. Our model not only outperformed publicly available tools such as Exomiser and LIRICAL, which are limited to SNV/INDEL prioritization, but also was able to prioritize a different type of variant, CNV, jointly with SNV/INDEL. When CNV, a different variant type that uses different variables than SNV/INDEL, was mixed into the variant list, our model successfully encoded all variants with high Bayesian score and symptom similarity score to be semantically clustered together on the manifold. Furthermore, our model can predict the genomic test results as positive or negative, which is not even an available function for the other publicly available tools.

The most important feature for both VP and predicting the genomic test results was the symptom similarity between the patient’ phenotypes and the disease phenotypes. This finding is in line with previous research that also suggested phenotype-driven approach to be effective in variant filtering and identification of the disease-causing variants^34^. To the best of our knowledge, the dataset that we used to evaluate our model is the largest real-world rare disease patient’s sequencing data that has been used for evaluation VP tools. Here, we used 4,646 of patients in the test dataset including 1,631 rare disease patients, and 3,015 of patients with negative test result among 23,115 of patients. Other VP tools had used either smaller real-world patient’s dataset or synthetic dataset due to challenges in collecting rare disease patient’s data, mainly because of privacy issues ^2,12,32,35,36^. Our extensive genomic dataset from real patients was exceptionally invaluable for this study as it was labelled with real clinical information.

Along this line, however, because we are relying on a real-world dataset, it is possible that there were false negative or positive test results within our dataset because of insufficient or incorrect clinical information. A variant/gene is selected for reporting only if the variant/gene can explain the provided phenotype of each patient. If the variant is within a gene/disease with later-onset, variable expressivity or incomplete penetrance, a disease-causing variant may not be reported because the symptoms do not match. It is also possible that the physician did not recognize a subset of the symptoms and missed to provide. Another possibility is an incorrect sample being sent to the laboratory for testing. Therefore, to minimize the risk of a sample swap, when the medical genetics team performs variant interpretation and encounters a pathogenic variant in an early-onset, full penetrance disease gene, they contact the physician to ask for more clinical information. However, even with these efforts, false negative or positive results could arise. Another variable that can introduce a bias into our modeling is that it was based on the customized ACMG variant interpretation guideline that a single reference laboratory developed. Because the overall diagnostic rate of this laboratory is comparable to others, they monitor the concordance of their ClinVar submission to other laboratories and they continuously receive feedback from the ordering physicians on their reports, we can assume that their customization is reasonable. However, further study is needed to see how the performance of our model changes when applied to other laboratories data and see if there is room for improvement, even though we expect that the noisiness is sufficiently reduced by using a large amount of data for training (n > 19,551).

In conclusion, we propose a model that uses multiple instance learning (MIL) and feed multiple inputs to predict the genomic test results and to jointly prioritize SNV/INDEL and CNV to assist the medical geneticist in determining which variant is most likely to be causal for the patient’s phenotype and eventually accelerate the variant interpretation process. This retrospective study is the first to show that deep learning-based VP can be improved with LTR for optimal prioritization.

## Data Availability

The model and experimental data are available at https://github.com/4pygmalion/ASC3.

## Acknowledgement

This work was supported by the Institute of Information & communications Technology Planning & Evaluation (IITP) grant funded by the Korea government (MSIT) (2022-0-00333, Multi-faceted analysis of pediatric rare disease Al integrated SW solution development)

