## Supplementary data for "A variant prioritization tool leveraging multiple instance learning for rare Mendelian disease genomic testing"

### Supplementary document 1

Supplementary table 1. Description of input features for SNV/INDEL.

| Feature name | Description | Source | Type | Range/Level |
| --- | --- | --- | --- | --- |
| acmg_bayesian | calculated posterior probability by Tavtigian, Sean V et al. | EVIDENCE | Float | [0, 1] |
| symptom_similarity | Calculated similarity between set of patient symptoms and set of disease phenotypes by Köhler et al. | EVIDENCE | Float | [0, 1] |
| qual | Phred-scaled probability that the site has no variant. | VCF | Integer | [0, $\infty$) |
| inhouse_af | Variant fraction in inhouse data | EVIDENCE | Float | [0, 1] |
| vaf | variant allele fraction | VCF | Float | [0, 1] |
| is_incomplete_zygosity | Mismatch between patient genotype and disease inheritance | EVIDENCE | Boolean | True/False |
| gnomad_gene_pLI | the probability of being loss-of-function intolerant score | gnomAD v2 | Float | [0, 1] |
| gnomad_gene_loeuf | loss-of-function observed/expected upper bound fraction | gnomAD v2 | Float | [0, 2] |
| SPLICEAI_VALUE | predicted effect of variant on splicing by Jaganathan et al. | spliceAI | Float | [0, 1] |
| wes_AC | gnomad allele count for WES | gnomAD v2 | Integer | [0, $\infty$) |
| wgs_AC | gnomad allele count for WGS | gnomAD v2 | Integer | [0, $\infty$) |
| clinvar_variant_scv_pathogenicity_n_p | Number of pathogenic records about variant in ClinVar | ClinVar | Integer | [0, $\infty$) |
| clinvar_variant_scv_pathogenicity_n_b | Number of benign records about variant in ClinVar | ClinVar | Integer | [0, $\infty$) |
| PVS1 | Encoded PVS1 rule on a scale of 0 to 5 with modifications. | ACMG/AMP guideline  EVIDENCE | Integer | 0/1/2/3/4/5 |
| PS1, 2, 3, 4 | Encoded PS rule on a scale of 0 to 5 with modifications. | ACMG/AMP guideline  EVIDENCE | Integer | 0/1/2/3/4/5 |
| PM1, 2, 3, 4, 5, 6 | Encoded PM rule on a scale of 0 to 5 with modifications. | ACMG/AMP guideline  EVIDENCE | Integer | 0/1/2/3/4/5 |
| PP1, 2, 3, 4, 5 | Encoded PP rule on a scale of 0 to 5 with modifications. | ACMG/AMP guideline  EVIDENCE | Integer | 0/1/2/3/4/5 |
| BA1 | Encoded BA rule on a scale of 0 to 5 with modifications. | ACMG/AMP guideline  EVIDENCE | Integer | 0/1/2/3/4/5 |
| BS1, 2, 3, 4 | Encoded BS rule on a scale of 0 to 5 with modifications. | ACMG/AMP guideline  EVIDENCE | Integer | 0/1/2/3/4/5 |
| BP1, 2, 3, 4, 5, 6, 7 | Encoded BP rule on a scale of 0 to 5 with modifications. | ACMG/AMP guideline  EVIDENCE | Integer | 0/1/2/3/4/5 |

Supplementary table 2. Description of input features for CNVs

| Feature name | Description | Source | Type | Range/Level |
| --- | --- | --- | --- | --- |
| acmg_bayesian | calculated posterior probability by Tavtigian, Sean V et al. | EVIDENCE | Float | [0, 1] |
| symptom_similarity | Maximal symptom similarity with diseases covered in genes [1] | EVIDENCE | Float | [0, 1] |
| gene_numbers | Number of affected by CNV | EVIDENCE | Int | [1, 1000) |

[1] Köhler, S., Schulz, M. H., Krawitz, P., Bauer, S., Dölken, S., Ott, C. E., Mundlos, C., Horn, D., Mundlos, S., & Robinson, P. N. (2009). Clinical Diagnostics in Human Genetics with Semantic Similarity Searches in Ontologies. American Journal of Human Genetics, 85(4), 457–464. <https://doi.org/10.1016/j.ajhg.2009.09.003>

[2] Tavtigian, S. V., Greenblatt, M. S., Harrison, S. M., Nussbaum, R. L., Prabhu, S. A., Boucher, K. M., Biesecker, L. G., & ClinGen Sequence Variant Interpretation Working Group (ClinGen SVI) (2018). Modeling the ACMG/AMP variant classification guidelines as a Bayesian classification framework. Genetics in medicine : official journal of the American College of Medical Genetics, 20(9), 1054–1060. https://doi.org/10.1038/gim.2017.210

### Supplementary document 2

Supplementary figure 1. Entire feature importance

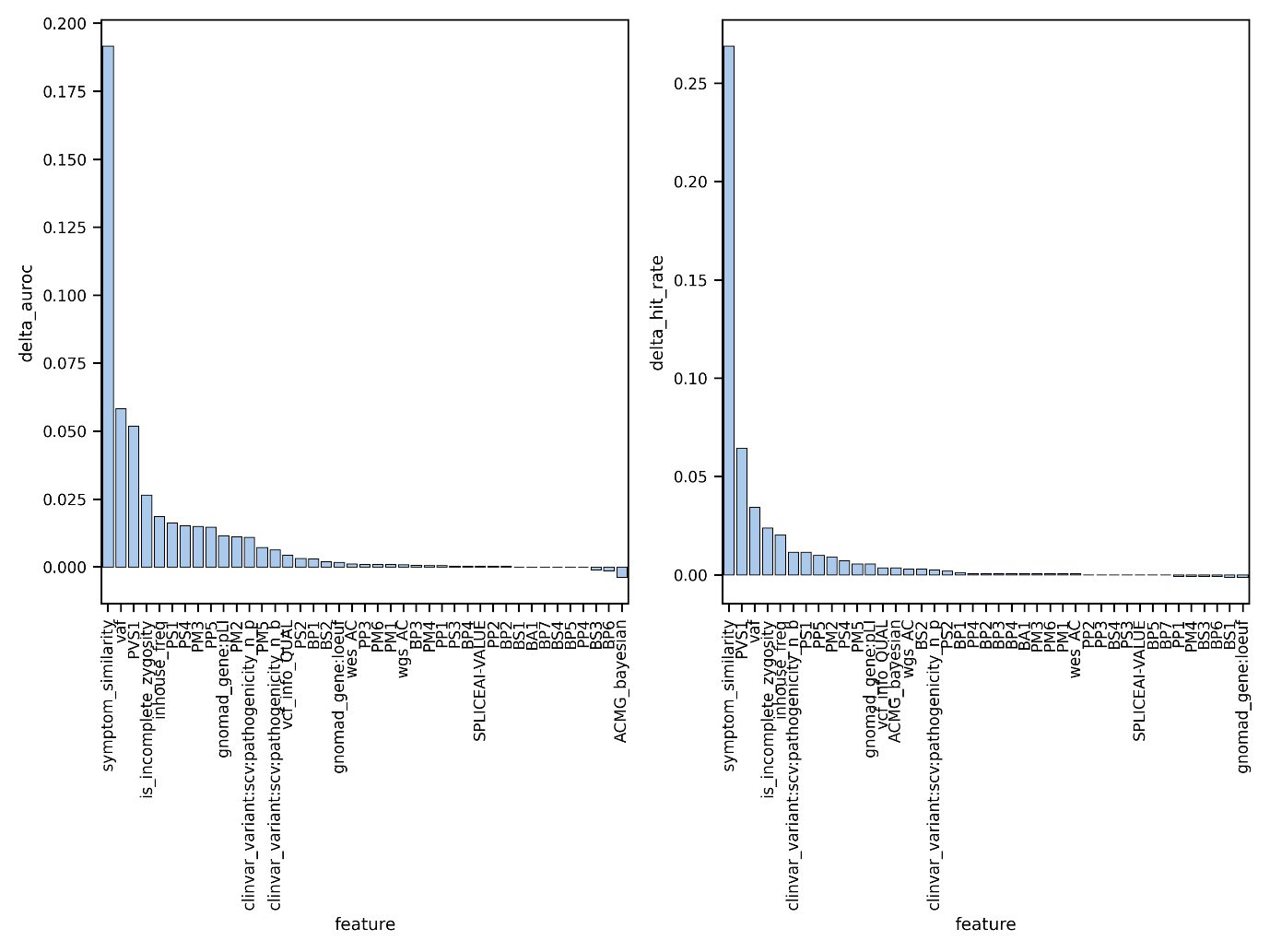

### Supplementary document 3

We trained our model with multi-task learning (MTL) for both prediction of genetic testing conclusion and variant prioritization. Generally, MTL improves the generalization performance of multiple learning task by sharing knowledge among those tasks ^1^. However, previous study empirically showed MTL model underperform on some task^2, 3^. This phenomenon is called the *negative sharing*. In our study, proposed model showed negative sharing in task for prioritization and prediction for genetic testing conclusion. We thought the negative sharing in our model by enforcing all the tasks will impair testing conclusion. To address this problem, Safe multi-task learning could be adopted for optimal performance.

Supplementary table 3. Entire ablation test

| Metrics | Models | | *P*-value |
| --- | --- | --- | --- |
|  | 3ASC  (pointwise RankNet) | 3ASC  (w/o pointwise RankNet) |  |
| Testing conclusion, F1, (mean, (SD)) | 0.831 (0.016) | 0.865 (0.007) | 0.007 |
| Hit rate @ 5 (SNVS/SMALL INDELS+CNV), (mean, (SD)) | 0.968 (0.003) | 0.942 (0.025) | 0.007 |
| Hit rate @ 5 (CNV), (mean, (SD)) | 0.942 (0.020) | 0.822 (0.039) | 0.007 |

[1] Yue, Z., Ye, F., Zhang, Y., Liang, C. & Tsang, I. W. Deep Safe Multi-Task Learning. (2021).

[2] Pengsheng Guo, Chen-YuLee, and Daniel Ulbricht. Learning to branch for multi-tasklearning (2020).

[3] Giwoong Lee, Eunho Yang, and SungHwang. Asymmetric multi task learning based on task relatedness and loss. InICML (2016)
